# Dissecting the Genetic Link Between Insulin Resistance and Coronary Heart Disease: A Multi-Trait GWAS and Proteomic-Metabolomic Mediation Analysis

**DOI:** 10.64898/2026.08.02.26359507

**Authors:** Rui Dong, Jiao Fu, Bowen Zhang, Qingling Zhang, Yang Yan, Guoliang Li

**Affiliations:** Hetao Institute of Mathematics and Interdisciplinary Sciences, Shenzhen-Hong Kong International Science Park 3 Binlang Road, Futian District, Shenzhen, 518000, P.R. China; Department of Endocrinology and Metabolism, the First Affiliated Hospital of Xi’an Jiaotong University, No. 277 Yanta West Road, Xi’an, 710061, P.R. China; Department of Cardiovascular Surgery, the First Affiliated Hospital of Xi’an Jiaotong University, No. 277 Yanta West Road, Xi’an, 710061, P.R. China; Tianjin Key Laboratory of Ionic-Molecular Function of Cardiovascular Disease, Department of Cardiology, Tianjin Institute of Cardiology, Second Hospital of Tianjin Medical University, Tianjin, China; Department of Cardiovascular Medicine, the First Affiliated Hospital of Xi’an Jiaotong University, 277 Yanta West Road, Xi’an China

**Keywords:** insulin resistance, coronary heart disease, polygenic risk score, serial mediation, plasma proteomics, HDL lipoprotein subclasses

## Abstract

Insulin resistance (IR) is a hallmark feature of Type 2 Diabetes Mellitus and a well-established risk factor for coronary heart disease (CHD), yet the pathways through which IR contributes to CHD remain incompletely understood. To comprehensively describe the genetic architecture of IR, we collected three direct IR indicators---fasting insulin (N = 151,013), the Modified Stumvoll insulin sensitivity index (N = 53,657), and pro-insulin (N = 45,861), and conducted joint analysis on them using Linkage Disequilibrium (LD) Score Regression and factor analysis. A single latent IR factor was generated which captures 60.5% of the shared genetic variance. Based on the IR factor, a multivariate GWAS was conducted within Genomic Structural Equation Modelling and then we built a polygenic risk score (PRS) of IR for the UK Biobank European cohort (N = 407,767; 55,729 CHD cases and 352,038 controls). Then we performed the association study to quantify the relationship between genetically predicted IR and CHD. Causal mediation analyses were also performed through 2,923 plasma proteins (Olink) and 168 metabolites (Nightingale NMR), followed by serial-mediation models (IR -> protein -> metabolite -> CHD) over all prescreened pairs. Single-mediator screening nominated 938 proteins and 159 metabolites; this yielded 3,669 significant "protein -> metabolite -> CHD" serial-mediation pathways, from which 33 broadly-acting core mediating proteins were prioritized. These acted predominantly through the large and very large high-density lipoprotein (HDL) particle subclasses: 30 of the 33 core proteins and all 14 HDL-subclass metabolites formed 306 pathways, of which 303 (99.0%) amplify the CHD risk. These results map a proteomic and metabolomic axis linking the genetic component of IR to CHD and highlight HDL lipoprotein-subclass biology as a candidate therapeutic and biomarker space.

## 1. Introduction

Coronary heart disease (CHD) is the leading cause of mortality worldwide and a major contributor to cardiovascular morbidity, accounting for an estimated nine million deaths annually^1^. Type 2 Diabetes Mellitus (T2DM) and its precursor states are well-established risk factors for CHD: individuals with diabetes carry approximately a two-fold elevated risk of a wide range of vascular diseases, independently from other conventional risk factors^2^. Despite this strong epidemiological association, the molecular pathways linking diabetes and its metabolic antecedents to coronary atherosclerosis are only partially understood, limiting our ability to identify high-risk individuals and to develop targeted preventive and therapeutic strategies.

Insulin resistance (IR), the hallmark physiological feature of T2DM, is a strong candidate mechanistic link between metabolic dysfunction and CHD. IR is characterized by impaired cellular responsiveness to insulin, leading to compensatory hyperinsulinemia and a cascade of downstream metabolic, inflammatory, and vascular sequelae. Observational and Mendelian-randomization studies have implicated IR in atherogenic dyslipidemia, endothelial dysfunction, low-grade systemic inflammation, and pro-thrombotic states --- each of which can plausibly contribute to coronary atherogenesis^3,4^. However, IR is a complex metabolic phenotype that cannot be captured by a single biomarker. Instead, it is represented by a heterogenous spectrum of related traits, including fasting insulin, insulin sensitivity indices (ISI), and pro-insulin levels, each capturing complementary aspects of IR and glucose homeostasis. A genetic study built on any one indicator therefore risks both attenuated effects and an incomplete picture of the biology these traits share. That shared biology, however, is precisely what can be recovered. Rather than forcing a choice among the individual indicators, their common genetic basis can be modelled jointly to yield a better-powered representation of IR. LD Score Regression (LDSC) uses GWAS summary statistics, rather than individual-level data, to estimate the SNP-based heritability of each trait and the genetic correlations among them^5^; because sample overlap between studies is absorbed into the LDSC intercept rather than the slope, the genetic-correlation estimates remain unbiased even when the contributing GWAS share participants^6^. Where the indicators prove strongly genetically correlated, this shared signal can be formalized in a structural model: Genomic Structural Equation Modelling (Genomic SEM) takes the genetic covariance matrix estimated by LDSC and fits a latent factor that captures the variance common to the indicators while separating it from trait-specific genetic effects^7^. By concentrating the genetic variance common to the indicators, this latent factor provides a representation of IR that is less dominated by trait-specific noise than any single measure, and the same framework yields multivariate GWAS summary statistics for the latent trait that feed directly into downstream polygenic scoring and mediation analysis.

Establishing a genetic signal, however, shows only that IR matters, not how it acts. Tracing how genetically higher IR raises CHD risk requires resolving the molecular intermediates that lie between them, the circulating proteins and metabolites through which genetic predisposition is ultimately translated into disease. Two features of the UK Biobank now make this tractable: large-scale plasma proteomics (Olink Explore, ∼3,000 proteins)^8^ and quantitative NMR metabolomics (Nightingale Health, 168 biomarkers spanning lipoprotein-particle subclasses, lipids, fatty acids, amino acids, ketone bodies, and inflammatory markers)^9^. These intermediates do not act in isolation: many are linked by directional biological dependencies, such as a protein regulating the level of a downstream metabolite, so the effect of IR may propagate through an ordered chain rather than through any single mediator. Testing one mediator at a time cannot recover such chains; a serial-mediation framework that models an explicit sequence (IR -> protein -> metabolite -> CHD) is needed to identify them.

These two ingredients, a robust genetic index of IR and a molecular account of how it reaches the coronary vasculature, have each been developed, but in isolation from one another. On the genetic side, multi-trait analyses have repeatedly outperformed single-trait GWAS for insulin resistance: early work combined fasting insulin with HDL cholesterol and triglycerides to sharpen locus discovery^10^, and more recent multivariate GWAS have assembled broader trait panels into composite IR phenotypes that are causally associated with multiple cardiometabolic diseases and that nominate candidate drug targets^11^. On the molecular side, the population-scale proteomic and metabolomic assays now available in the UK Biobank have been used both to improve prediction of incident cardiovascular disease^12,13^ and to identify individual plasma proteins that mediate the progression from metabolic conditions such as diabetes to downstream cardiovascular outcomes^14^. Three features, however, limit how far this mediation work can illuminate the IR–CHD route. First, the exposure is typically a clinically diagnosed condition rather than genetic liability, leaving estimates vulnerable to reverse causation and to confounding by shared risk factors. Second, mediators are examined one molecular layer at a time, with each protein or metabolite treated as an independent conduit, even though the intermediates are biochemically coupled in the ordered fashion described above. Third, the emphasis has fallen on predictive performance or on cataloguing individual associations rather than on reconstructing the route from cause to disease. As a consequence, the specific chain by which genetically elevated IR propagates through the circulating proteome and metabolome to raise CHD risk has not been mapped, and it is this gap that the present study addresses.

In this study, we integrated multi-trait genetic discovery with proteomic and metabolomic serial mediation to resolve the molecular path from IR to CHD. Using European-ancestry GWAS summary statistics that exclude UK Biobank to avoid overlap with our target cohort, we first characterized the genetic architecture of three IR-related metabolic traits, fasting insulin, ISI, and pro-insulin, estimating SNP-based heritability and pairwise genetic correlations, testing their suitability for a single-factor representation, and conducting a multivariate GWAS of the resulting latent IR factor in Genomic SEM. We then constructed a Bayesian polygenic risk score (PRS) for the latent factor using PRS-CS^15^ and apply it in the UK Biobank European cohort, where genetic IR, plasma proteins, circulating metabolites, and incident CHD are measured. Within this cohort we screen 2,923 proteins and 168 metabolites as candidate mediators, then fit a serial-mediation model (IR PRS -> protein -> metabolite -> CHD) across the prescreened protein–metabolite pairs to identify the dominant pathways. In contrast to prior work, which has typically relied on single IR proxies and single-mediator tests, this design combines a latent multi-trait IR factor with genome-to-outcome serial mediation across thousands of molecular intermediates, moving from genetic susceptibility to IR toward interpretable, potentially actionable molecular intermediaries of CHD risk.

## 2. Methods

### 2.1 Discovery GWAS summary statistics and quality control

We analysed three complementary indicators of insulin resistance: fasting insulin, the Modified Stumvoll insulin sensitivity index (ISI), and pro-insulin. For each trait we used the largest publicly available European-ancestry GWAS that did not include UK Biobank participants, i.e., fasting insulin (N = 151,013)^16^, the Modified Stumvoll ISI (N = 53,657)^17^, and pro-insulin (N = 45,861)^18^, so as to avoid sample overlap with the target cohort used in all downstream analyses. All three sets of summary statistics were reported on genome build GRCh37, were derived from BMI-adjusted association models at source, and were drawn from the European-ancestry stratum of each study, with no UK Biobank participants. Using BMI-adjusted statistics indexes insulin resistance conditional on adiposity rather than the correlated variation attributable to overall obesity. Quality control was performed with EasyQC (v23.8)^19^ using the 1000 Genomes Phase 3 European panel^20^ as the reference. Variants were processed in two passes. In the first pass, we applied SNP-level filters, removing variants that met any of the following criteria: alleles other than A, T, C, or G; monomorphic sites; minor allele frequency (MAF) below 0.5% or missing MAF; duplicated or multiallelic variants; and, where imputation quality was reported, an imputation INFO score below 0.9^21^. In the second pass, the surviving variants were aligned to the 1000 Genomes Phase 3 European reference by chromosomal position, and effect and other alleles were checked against the reference to resolve strand ambiguities; variants that failed reference-allele matching, had a MAF below 0.5%, or lacked a p-value were excluded. Remaining variants were annotated with rsIDs using gwaslab (v3.5.8)^22^ against the 1000 Genomes dbSNP151 reference (hg19/GRCh37)^23^ and the NCBI RefSeq GCF_000001405.25 reference genome^24^.

### 2.2 SNP-based heritability and genetic correlations

Before combining the three indicators, we characterised their genetic architecture using LD Score Regression (LDSC). The LD reference panel is also the 1000 Genomes Phase 3 European panel^20^ to match the ancestry of the summary statistics. SNP-based heritabilities and pairwise genetic correlations were estimated on the observed scale using the multivariable LDSC implementation in GenomicSEM (v0.0.5)^7^ in R (v4.4.3), which jointly estimates the genetic covariance matrix across the three traits and its associated sampling covariance matrix from the munged summary statistics. Estimating these two matrices together propagates the uncertainty in, and the dependencies among, the trait-level estimates, including any induced by sample overlap between the contributing GWAS, into all subsequent modelling steps.

### 2.3 Latent factor model and multivariate GWAS

Using the genetic covariance matrix and its sampling covariance matrix estimated above, we specified a common-factor structural equation model in which a single latent factor (F1) was defined by the three IR indicators, with F1 taken to represent the genetic liability to insulin resistance shared across fasting insulin, the Modified Stumvoll ISI, and pro-insulin. The model was fit by diagonally weighted least squares (DWLS), the default estimator in GenomicSEM. Because a single-factor model of three indicators is just-identified (zero degrees of freedom), conventional global fit indices are uninformative; single-factor adequacy was therefore assessed by a complementary exploratory factor analysis of the genetic covariance matrix, quantifying the proportion of genetic variance explained by a single factor, together with the pattern and magnitude of the standardized factor loadings. Consistent with the ISI being an index of insulin sensitivity rather than resistance, it was expected to load on F1 with sign opposite to fasting insulin and pro-insulin at this stage.

For the multivariate genome-wide association step, the full post-QC summary statistics (Section 2.1) were aligned to the 1000 Genomes Phase 3 European reference panel, excluding insertions/deletions and variants with imputation quality below 0.6; 7,880,834 SNPs remained for analysis. Prior to this step, the sign of the ISI effect estimates was reversed so that all three indicators were oriented in a common direction, with higher values corresponding to greater insulin resistance; F1 was thereby scaled so that higher genome-wide F1 values index greater insulin resistance. At each SNP we estimated the association with F1 under the common-factor model. GenomicSEM additionally returned the per-SNP heterogeneity statistic Q_SNP_, which tests whether a SNP’s effects across the three indicators are consistent with an effect operating through the common factor rather than through indicator-specific pathways. Independent genome-wide-significant loci, for both the F1 association and Q_SNP_ (P < 5×10), were defined by LD-based clumping in PLINK (index P < 5×10 ; r² < 0.1 within ±1 Mb) against the same 1000 Genomes Phase 3 European reference panel.

### 2.4 Polygenic risk score construction

A polygenic risk score for insulin resistance was constructed from the multivariate IR-factor summary statistics using PRS-CS^25^ with the 1000 Genomes Phase 3 European LD reference panel. Because F1 was oriented so that higher values index greater insulin resistance (Section 2.3), higher scores likewise correspond to greater genetic liability to insulin resistance. Only variants shared between the F1 summary statistics and the UK Biobank imputation panel were retained. Posterior SNP effect sizes were estimated under the Bayesian continuous shrinkage prior with a fixed global shrinkage parameter (φ = 0.01) and the package’s default scale and shape hyperparameters (a = 1, b = 0.5); the Gibbs sampler was run for 1,000 iterations with the first 500 discarded as burn-in and a thinning interval of 5, on each of the 22 autosomes in parallel.

### 2.5 UK Biobank cohort and phenotypes

All downstream polygenic scoring, proteomic, metabolomic, and mediation analyses were conducted in the UK Biobank, restricted to a quality-controlled European-ancestry cohort of 407,767 individuals. Sample-level quality control followed standard UK Biobank practice: restriction to genetically inferred European ancestry, and exclusion of individuals with sex-chromosome aneuploidy or a mismatch between genetic and reported sex, excess heterozygosity or high genotype missingness, and participants who had withdrawn consent.

The primary outcome was coronary heart disease (CHD), ascertained by combining self-reported, hospital inpatient, and algorithmically defined records. Self-reported cases were identified from Field 20002 (codes 1074/1075) and Field 6150 (codes 1/2); hospital inpatient cases from International Classification of Diseases (ICD)-10 codes I20– I25 and ICD-9 codes 410–414 (Fields 41270/41202/41204 and 41271/41203/41205); and algorithmically defined cases from the first-occurrence records for I20–I25 (Fields 131296–131306). Both prevalent and incident cases were retained, yielding 55,729 CHD cases and 352,038 controls in the quality-controlled European cohort.

Individual-level polygenic risk scores were computed in the UK Biobank imputed genotype data for the quality-controlled European participants. For each individual, the retained variants (Section 2.4) were weighted by their PRS-CS posterior effect sizes and summed across the 22 autosomes to yield a single insulin-resistance polygenic risk score, oriented so that higher scores index greater genetic liability to insulin resistance

### 2.6 Plasma proteomic and metabolomic mediators and covariate adjustment

Candidate mediators were drawn from two orthogonal molecular layers assayed in UK Biobank participants. Plasma proteins were obtained from the UK Biobank Pharma Proteomics Project (UKB-PPP)^8^, which measured 2,923 proteins on the Olink Explore platform in approximately 50,000 participants; protein abundances are expressed as normalised protein expression (NPX) values. Circulating metabolites were obtained from the UK Biobank Nightingale Health nuclear magnetic resonance (NMR) platform^9^, which quantifies 168 biomarkers in approximately 500,000 participants. These biomarkers are dominated by lipoprotein-subclass measures, capturing lipid concentration and composition across 14 subclasses, alongside routine lipids, fatty acids and fatty-acid composition, amino acids, ketone bodies, glycolysis-related metabolites, and the inflammation marker glycoprotein acetyls. Only baseline-visit measurements were retained, and the biomarker values were processed to remove technical variation, including sample-preparation time, spectrometer batch and drift, and well-position effects, and to flag outliers using the ukbnmr R package^26^.

All mediation models adjusted for a common set of covariates capturing demographic, socioeconomic, and lifestyle sources of confounding: age at recruitment, sex, the first ten genetic principal components, the Townsend deprivation index, highest educational qualification, employment status, physical activity, smoking status, and self-reported sleeplessness. Because a mediator and the outcome may share common causes, adjusting only the exposure is insufficient: unadjusted confounding of the mediator–outcome and exposure–outcome relationships would bias the estimated indirect effects. We therefore applied this covariate set throughout the mediation models (Section 2.7), rather than to the exposure alone, so that each estimated path was adjusted for the same measured confounders.

### 2.7 Mediation analysis

Mediation analyses proceeded in two stages: a single-mediator prescreening of proteins and metabolites, followed by serial mediation modelling of the prescreened pairs. Throughout, the exposure was the insulin-resistance polygenic risk score (Section 2.4) and the outcome was CHD case status (Section 2.5). Causal interpretation of the estimated indirect effects rests on the usual sequential-ignorability assumptions, no unmeasured exposure–mediator, exposure–outcome, or mediator–outcome confounding, addressed here by adjusting every path for the covariate set defined in Section 2.6.

#### Single-mediator prescreen

Each candidate mediator was tested individually. Plasma proteins levels were analyzed as normalized protein expression (NPX) levels, which the UKB-PPP provides on a log_2_ scale and therefore require no further transformation. For each plasma protein, a causal-mediation analysis was conducted following the framework^27^, with the IR polygenic risk score as the exposure, the protein as the mediator, and CHD as the outcome. Average causal mediation (indirect) effects and total effects were estimated, and the proportion mediated together with its bootstrap p-value were computed from 1,000 nonparametric bootstrap resamples. The identical procedure was applied separately to each of the 168 NMR metabolites. To ensure stable estimation, mediators were retained only where the complete-case sample size exceeded 30,000 for proteins, and 350,000 for metabolites, reflecting the coverage of the Olink and Nightingale assays respectively. Mediators whose proportion-mediated bootstrap p-values fell below 0.05 were carried forward, yielding 938 candidate proteins and 159 candidate metabolites.

#### Serial mediation

For every (protein, metabolite) pair obtained from the two prescreened sets, a structural equation model was fit using the lavaan package^28^ following the path structure PRS -> protein -> metabolite -> CHD. Proteins were placed upstream of metabolites on biological grounds: plasma proteins such as enzymes, transporters, and apolipoproteins act as regulators of metabolite generation, interconversion, and clearance, so that protein-level changes are mechanistically positioned to drive downstream shifts in metabolite concentrations rather than the reverse. Prior to fitting, both the protein and metabolite mediators were rank-based inverse-normal transformed and then, together with the IR PRS, residualized on the covariates listed in Section 2.6; CHD was retained as a binary outcome. The quantity of primary interest was the serial indirect effect (PRS -> protein -> metabolite -> CHD) and its proportion of the total effect, estimated alongside the two single-mediator indirect effects (via the protein alone, and via the metabolite alone).

Because the number of candidate protein–metabolite pairs was large (938 × 159 = 149,142), the full grid was evaluated with a two-pass scheme that balanced computational cost against statistical rigor. In Pass 1, every pair was fit once by maximum likelihood, with the standard error of the serial indirect effect approximated analytically by the delta method. Pairs reaching a serial-indirect p < 0.05 in Pass 1 advanced to Pass 2, in which the same model was refit with 1,000 nonparametric bootstrap resamples. Of the 149,142 candidate pairs, 115,645 pairs (77.5%) both passed the Pass 1 screen and converged in the bootstrap, forming the analysed set; the remaining pairs were either screened out in Pass 1 or failed to converge. Benjamini–Hochberg false-discovery-rate correction^29^ was then applied across the 115,645 converged pairsseparately for the serial indirect effect and for each of the two single-mediator indirect effects.

### 2.8 Ethics statement

This research was conducted using the UK Biobank Resource under Application Number 152047. UK Biobank holds approval from the North West Multi-centre Research Ethics Committee as a Research Tissue Bank (reference 21/NW/0157), and all participants provided written informed consent at recruitment. Analyses were performed on de-identified data in accordance with UK Biobank’s terms of access and governance framework. The three input GWAS summary statistics were obtained from previously published studies, each conducted with appropriate ethical approval and participant consent at source, and contained no individual-level or identifiable information. Because this work is a secondary analysis of existing de-identified data, no additional ethical approval was required. No animal experiments were performed.

## 3. Results

### 3.1 Genetic architecture of the three IR traits

We first characterized the genetic architecture of the three IR indicators and tested whether they support a single latent factor. After QC, 9,135,760, 9,336,961, and 9,404,237 autosomal variants remained for fasting insulin, ISI, and pro-insulin, respectively. SNP-based heritability on the observed scale (h², SE) was 0.094 (0.009) for fasting insulin, 0.109 (0.012) for the ISI, and 0.108 (0.022) for pro-insulin. Multivariable LD score Regression in GenomicSEM a pairwise genetic correlation matrix showing a strong inverse correlation between fasting insulin and ISI (r_g_ = −0.835), a moderate positive correlation between fasting insulin and pro-insulin (r_g_ = +0.345), and a weak-to-moderate inverse correlation between ISI and pro-insulin (r_g_ = −0.230) (Figure 1). LDSC intercepts indicated that the cross-trait sample overlap was the greatest between fasting insulin and the ISI cohorts (intercept = −0.199, SE 0.007), and the minimal between the fasting insulin and pro-insulin cohorts (intercept = 0.011, SE 0.006), consistent with the design of the contributing GWAS.A single common-factor model defined the latent IR factor (F1), which loaded strongly on fasting insulin (standardized loading 1.12, marginally exceeding 1.0 --- a minor Heywood case) and the ISI (−0.75) and more weakly on pro-insulin (0.31); a complementary exploratory factor analysis supported a single factor explaining 60.5% of the genetic variance across the three traits. F1 was carried forward as the basis for the multivariate GWAS.

**Figure 1.**
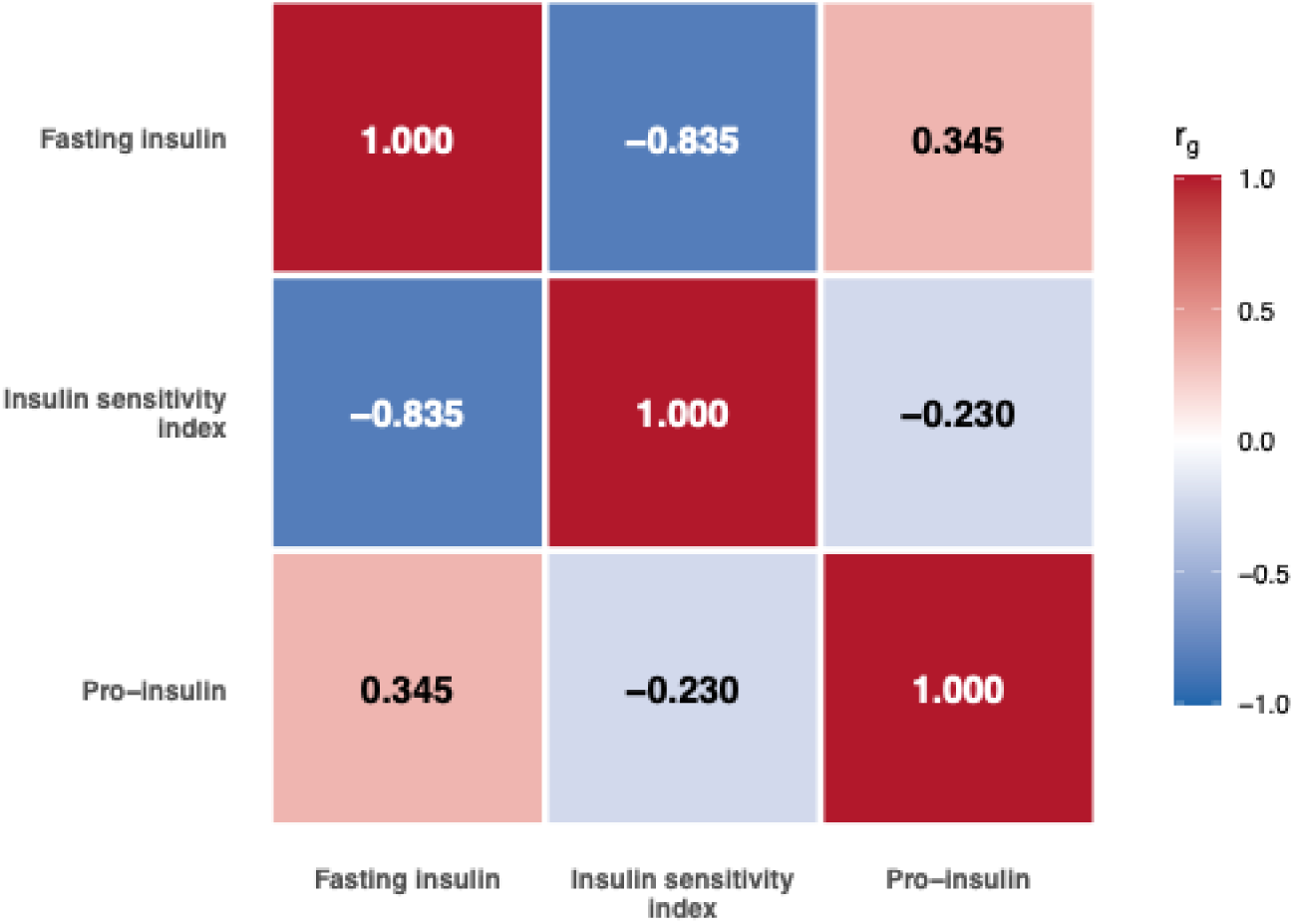
Genetic correlation among the three insulin-resistance traits. Pairwise genetic correlations (r_g_) estimated by multivariate LD-score regression (GenomicSEM) for fasting insulin, the insulin sensitivity index, and pro-insulin. Cell color encodes r_g_; hue indicates direction (blue, negative; red, positive) and intensity encodes magnitude on a fixed −1 to +1 scale.

### 3.2 Multivariate GWAS of the latent IR factor

The multivariate GWAS of the latent IR factor (F1) was fit at 7,880,834 SNPs shared across the summary statistics for the three traits. At the genome-wide-significance threshold (P < 5×10^-8^), 2,537 SNPs were significant for the F1 association, resolving into 74 independent loci by LD-clumping (Figure 2; Supplementary Table S1). The quantile– quantile plot showed only modest inflation (λ_GC_ = 1.16; Figure S1), consistent with polygenic signal rather than substantial confounding. Per-SNP heterogeneity across the three IR traits (Q_SNP_) reached genome-wide significance for 3,555 SNPs (86 independent loci), identifying variants whose effects diverge from the common-factor structure (Supplementary Table S2). The discovery sample size used for polygenic scoring was 63,743, the harmonic mean of the three contributing GWAS sample sizes.

**Figure 2.**
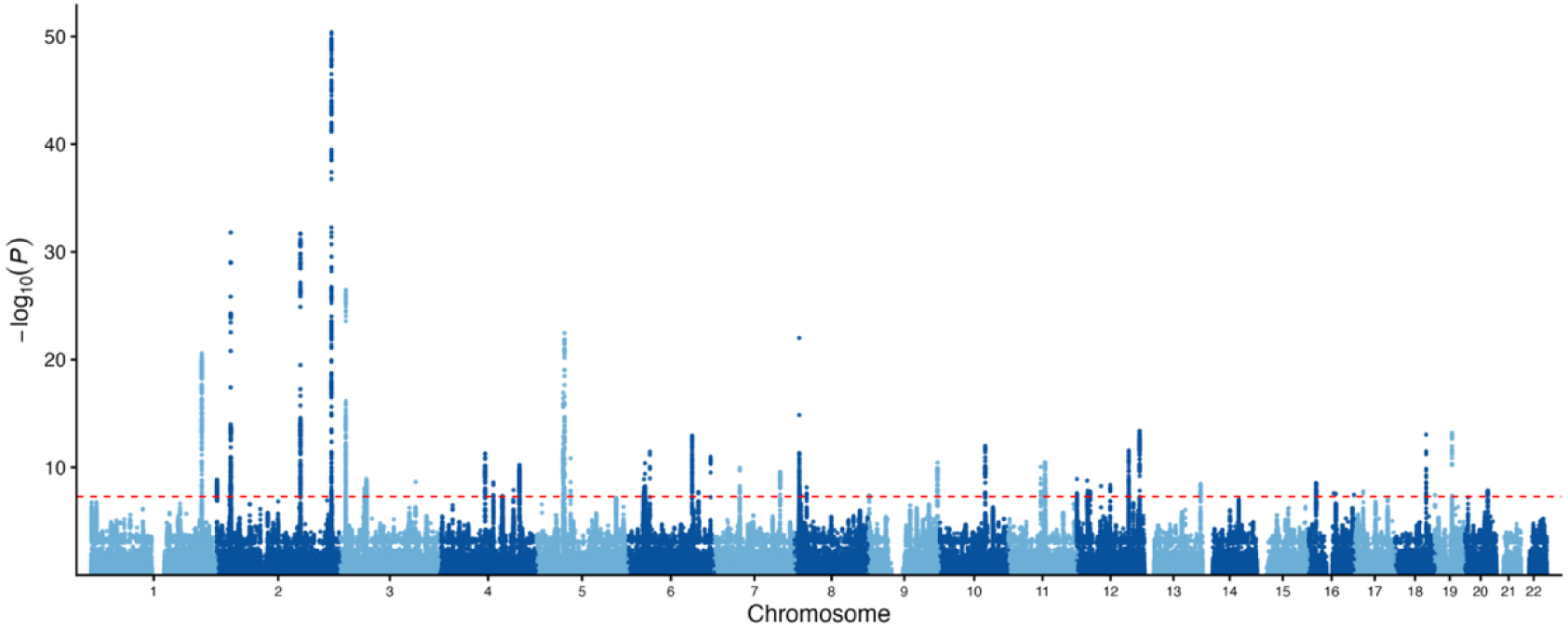
Manhattan plot of the multivariate GWAS of the latent insulin-resistance factor (F1). Each point is one of the 7,880,834 SNPs tested for association with the common factor F1, ordered by chromosomal position; the y-axis is −log (P) for the SNP–F1 association and the dashed line marks the genome-wide-significance threshold (P = 5×10^-8^).

### 3.3 Study cohort characteristics

All downstream analyses were conducted in the QC-passed European-ancestry UK Biobank cohort (N = 407,767), of whom 55,729 (13.7%) were CHD cases and 352,038 were controls. Olink Explore proteomic measurements (2,923 plasma proteins) were available for 43,658 participants, and Nightingale NMR metabolomic measurements (168 biomarkers, baseline visit) for 402,844 participants. Baseline demographic, lifestyle, and clinical characteristics of the cohort, stratified by CHD case status, are summarized in Table 1. Compared with controls, CHD cases were older (mean 61.0 vs 56.3 years), more often male (65.1% vs 42.9%), and had higher BMI (28.9 vs 27.2 kg/m²) and a markedly higher prevalence of type 2 diabetes (18.5% vs 6.4%), hypertension (79.0% vs 36.3%), and hyperlipidemia (70.6% vs 23.1%; all P < 0.001).

**Table 1.** Baseline characteristics of the study cohort by CHD status. Demographic, lifestyle, and clinical characteristics of the QC-passed European UK Biobank cohort (N = 407,767), stratified by CHD status. Continuous variables are summarized as mean (SD) and compared with a two-sample t-test; categorical variables as n (%) and compared with a chi-square test.

| Variable | Level | Overall (N=407,767) | Control (n=352,038) | Case (n=55,729) | P |
| --- | --- | --- | --- | --- | --- |
| Age at recruitment (years), mean (SD) |  | 56.9 (8.0) | 56.3 (8.0) | 61.0 (6.5) | <1e-300 |
| BMI (kg/m<sup>2</sup>), mean (SD) |  | 27.4 (4.8) | 27.2 (4.7) | 28.9 (5.0) | <1e-300 |
| Townsend deprivation index, mean (SD) |  | -1.6 (2.9) | -1.6 (2.9) | -1.1 (3.2) | 3.7e-296 |
| Physical activity (MET-min/wk), mean (SD) |  | 2667.8 (2661.6) | 2666.9 (2642.9) | 2673.7 (2782.4) | 0.638 |
| IR PRS, mean (SD) |  | -1.1 (0.8) | -1.1 (0.8) | -1.1 (0.8) | 1.9e-25 |
| Sex, n (%) | Female | 220,420 (54.1%) | 200,963 (57.1%) | 19,457 (34.9%) | <1e-300 |
|  | Male | 187,347 (45.9%) | 151,075 (42.9%) | 36,272 (65.1%) |  |
| **Smoking status, n (%)** | Current | 41,166 (10.1%) | 33,615 (9.5%) | 7,551 (13.5%) | <1e-300 |
|  | Never | 221,809 (54.4%) | 198,550 (56.4%) | 23,259 (41.7%) |  |
|  | Prefer not to answer | 1,429 (0.4%) | 1,118 (0.3%) | 311 (0.6%) |  |
|  | Previous | 143,363 (35.2%) | 118,755 (33.7%) | 24,608 (44.2%) |  |
| **Alcohol intake frequency, n (%)** | Daily or almost daily | 86,270 (21.2%) | 74,503 (21.2%) | 11,767 (21.1%) | 1.9e-280 |
|  | Never | 26,805 (6.6%) | 21,576 (6.1%) | 5,229 (9.4%) |  |
|  | Once or twice a week | 107,762 (26.4%) | 93,793 (26.6%) | 13,969 (25.1%) |  |
|  | One to three times a month | 45,209 (11.1%) | 39,401 (11.2%) | 5,808 (10.4%) |  |
|  | Prefer not to answer | 280 (0.1%) | 222 (0.1%) | 58 (0.1%) |  |
|  | Special occasions only | 43,191 (10.6%) | 36,167 (10.3%) | 7,024 (12.6%) |  |
|  | Three or four times a week | 98,250 (24.1%) | 86,376 (24.5%) | 11,874 (21.3%) |  |
| **Highest education, n (%)** | A levels/AS levels or equivalent | 46,491 (11.4%) | 41,617 (11.8%) | 4,874 (8.7%) | <1e-300 |
|  | College/University degree | 128,869 (31.6%) | 116,575 (33.1%) | 12,294 (22.1%) |  |
|  | CSEs or equivalent | 21,999 (5.4%) | 19,676 (5.6%) | 2,323 (4.2%) |  |
|  | None of the above | 71,017 (17.4%) | 54,735 (15.5%) | 16,282 (29.2%) |  |
|  | NVQ/HND/HNC or equivalent | 26,455 (6.5%) | 21,520 (6.1%) | 4,935 (8.9%) |  |
|  | O levels/GCSEs or equivalent | 90,683 (22.2%) | 79,499 (22.6%) | 11,184 (20.1%) |  |
|  | Other professional (e.g., nursing, teaching) | 19,037 (4.7%) | 15,872 (4.5%) | 3,165 (5.7%) |  |
|  | Prefer not to answer | 3,216 (0.8%) | 2,544 (0.7%) | 672 (1.2%) |  |
| **Type 2 diabetes, n (%)** |  | 32,969 (8.1%) | 22,665 (6.4%) | 10,304 (18.5%) | <1e-300 |
| **Hypertension, n (%)** |  | 171,948 (42.2%) | 127,923 (36.3%) | 44,025 (79.0%) | <1e-300 |
| **Hyperlipidemia, n (%)** |  | 120,723 (29.6%) | 81,393 (23.1%) | 39,330 (70.6%) | <1e-300 |

### 3.4 IR polygenic score and its association with CHD

The IR PRS, derived from the multivariate IR-factor summary statistics, was computed for all 407,767 participants from the 1,073,856 variants shared with the UK Biobank imputation panel. The resulting per-individual score was approximately normally distributed (mean = −1.11, SD = 0.75, median = −1.11; Figure 3). A higher IR PRS was associated with greater odds of CHD (OR = 1.05 per standard deviation, 95% CI 1.04-1.06; P = 1.3×10^-27^; logistic regression adjusted for age, sex, and ten genetic principal components).

**Figure 3.**
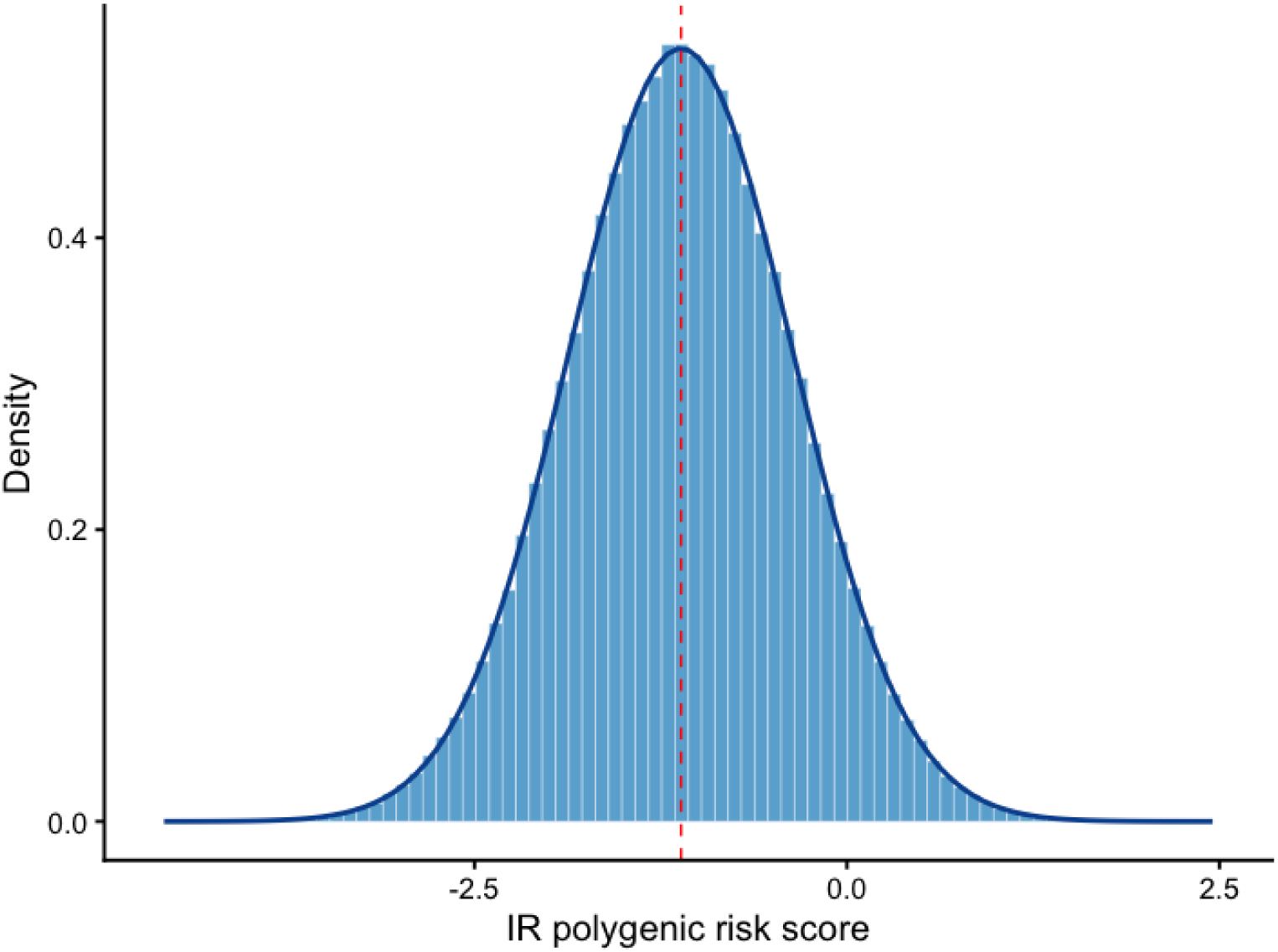
Distribution of the insulin-resistance polygenic risk score. Per-individual IR PRS for the 407,767 QC-passed European-ancestry UK Biobank participants. The histogram shows the empirical density; the blue curve is a normal density with the observed mean and SD, and the dashed line marks the mean (mean = −1.11, SD = 0.75, median = −1.11).

### 3.5 Single-mediator screen of candidate protein and metabolite mediators

To identify molecular intermediates of the IR->CHD relationship, each of the 2,923 Olink plasma proteins and 16 Nightingale NMR metabolites was tested individually as a candidate mediator between the IR PRS (exposure) and incident/prevalent CHD (outcome) using single-mediator causal mediation analysis (Methods Section 2.7). At a nominal bootstrap proportion-mediated *p* < 0.05, 938 of 2,923 proteins (32.1%) and 159 of 168 metabolites (94.6%) showed significant mediation and were carried forward as candidate single-step mediators (Supplementary Tables S3–S4).For both molecular layers the mediated effect was predominantly risk-increasing (837 of 938 proteins; 101 of 159 metabolites) --- higher genetic IR was associated with a mediator change that in turn tracked higher CHD risk ---, with a smaller set of protective signals (101 proteins, 58 metabolites).

Restricting to the strongest mediators (|proportion mediated| > 0.30), the leading protein mediators were established insulin-resistance and lipid-handling proteins, headed by adiponectin (ADIPOQ; proportion mediated 57%), followed by PRAP1, apolipoprotein F (APOF), PON3, galectin-4 (LGALS4), carboxypeptidase M (CPM), and hepatocyte growth factor (HGF), with N-terminal pro-B-type natriuretic peptide (NT-proBNP) the leading protective mediator (Table 2a). For metabolites, every mediator exceeding this threshold was an HDL-particle measure: the cholesterol, cholesteryl-ester, particle concentration, and size of large HDL (L-HDL) and very large HDL (XL-HDL) (Table 2b). This localizes the lipid signal to HDL particle biology, which we decompose by serial mediation below.

**Table 2.**
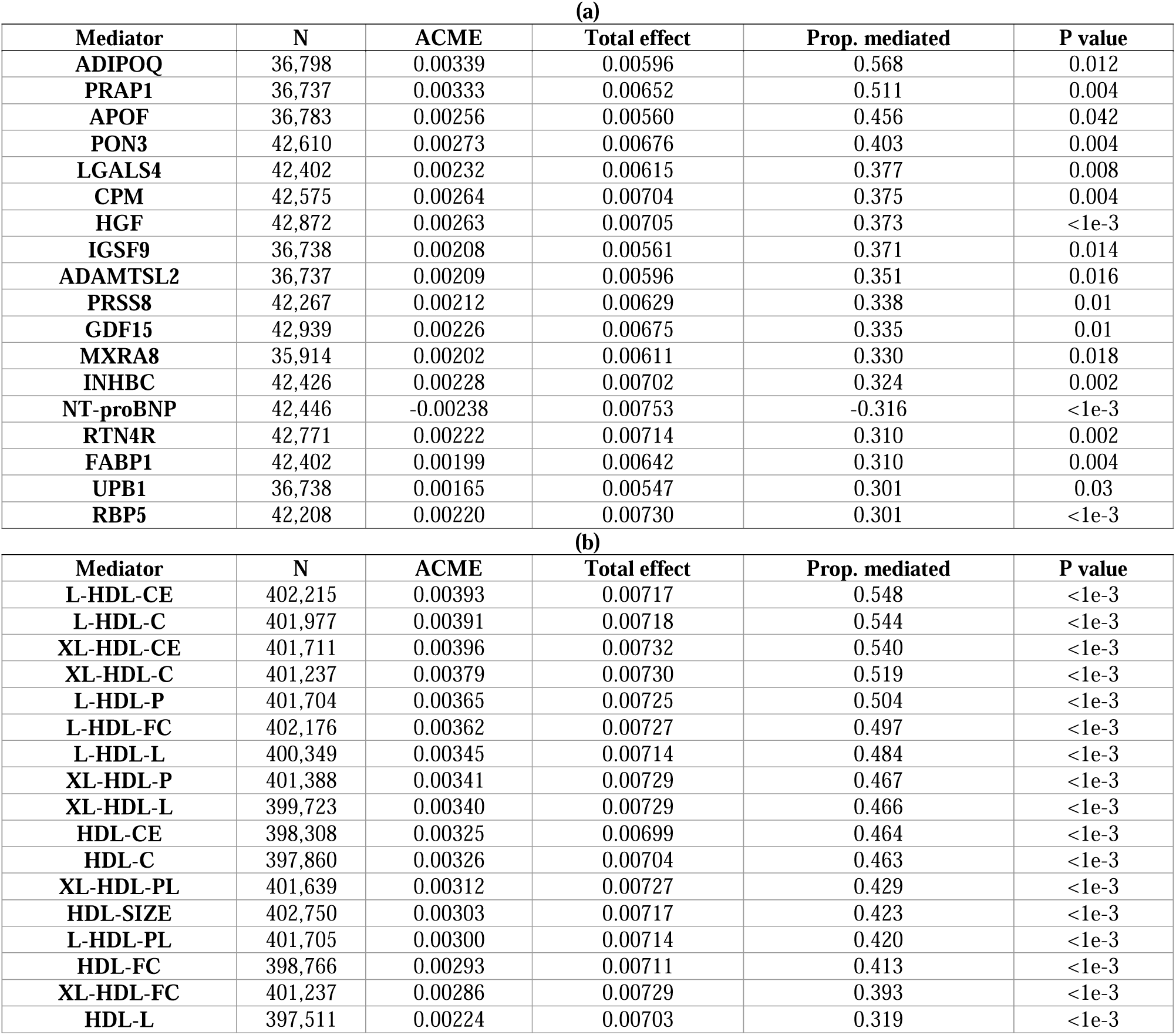
Top single mediators of the IR-PRS -> CHD effect. Single-step mediators most strongly transmitting the genetic IR effect to CHD, defined as a bootstrap proportion-mediated p < 0.05 and |proportion mediated| > 0.30. (a) plasma proteins (18); (b) NMR metabolites (17). Prop. Mediated stands for proportion of mediated effect among total effect. Lipoprotein-measure suffixes (Nightingale NMR): C, total cholesterol; CE, cholesteryl esters; FC, free cholesterol; PL, phospholipids; P, particle concentration; L, total lipids; SIZE, mean particle diameter. Subclass prefixes: XL, very large; L, large; M, medium; S, small.

### 3.6 Serial-mediation core proteins

To capture the sequential protein->metabolite dependencies that single-mediator analysis cannot see, we fit a serial-mediation structural-equation model (IR PRS -> protein -> metabolite -> CHD) to all 149,142 candidate triads formed by the 938 prescreened proteins and 159 prescreened metabolites (Methods Section 2.7). Reflecting the power afforded by the cohort, the serial indirect effect was significant for the large majority of triads --- 113,917 of the 115,645 that converged under bootstrapping (98.5%) at FDR < 0.05, and 115,481 (99.9%) with a 95% bootstrap confidence interval excluding zero. Because statistical significance was therefore minimally discriminating, we prioritized pathways by effect size, retaining those whose FDR-significant serial indirect effect explained > 5% of the total IR-PRS->CHD effect and whose protein-only and metabolite-only indirect effects were both FDR-significant. This left 3,669 pathways spanning 161 proteins and 134 metabolites (Supplementary Table S5), comprising 1,677 amplifying pathways --- in which higher genetic IR drives a protein->metabolite change associated with higher CHD risk (positive proportion mediated) --- and 1,992 compensatory pathways, in which the chain runs counter to the total effect (negative proportion mediated), consistent with homeostatic buffering.To distinguish broadly-acting mediators from narrowly-acting ones, we ranked the 161 proteins by the breadth and magnitude of their pathways (the number of significant pathways, the number of distinct metabolite classes spanned, and the maximum and mean absolute proportion mediated) and retained the 33 "core" proteins exceeding the 75th percentile on all four metrics (Figure 4; Table 3). Classifying each protein by the dominant direction of its pathways --- amplifying- or compensatory-dominant when > 70% of its pathways shared one sign, otherwise mixed --- gave six amplifying-dominant, 15 compensatory-dominant, and 12 mixed proteins (Figure 4). The amplifying-dominant set was an apolipoprotein cluster --- APOA1, APOM, APOD, and APOF --- together with MXRA8 and RTN4R; the compensatory-dominant set included LDLR, LCAT, IGFBP2, and GHR; and metabolically central proteins such as adiponectin (ADIPOQ), lipoprotein lipase (LPL), and PLTP showed mixed direction. These core proteins acted overwhelmingly through HDL lipoprotein-particle subclasses, which we examine next.

**Figure 4.**
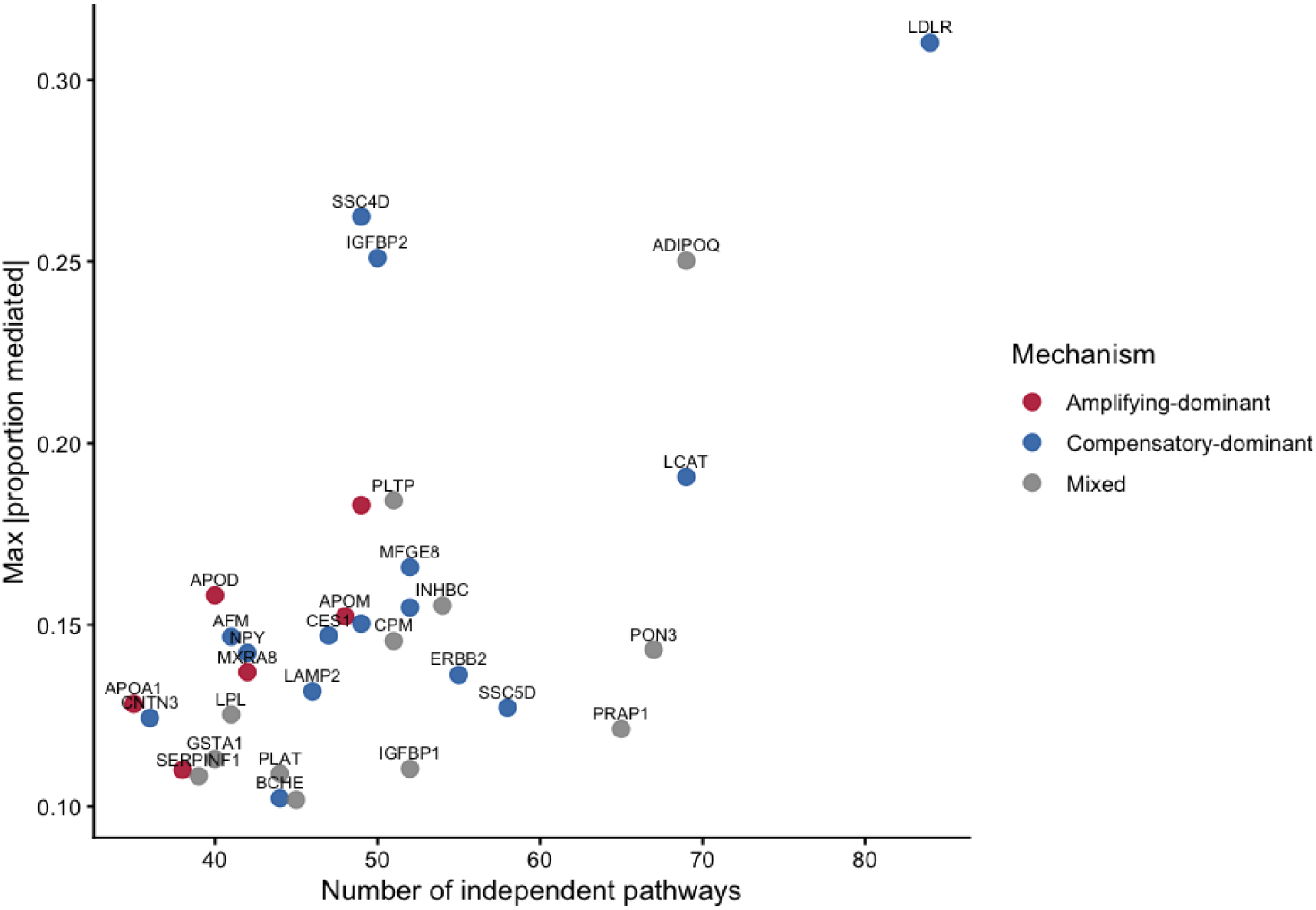
Landscape of the 33 core mediating proteins. Each point is one of the 33 broadly-acting core proteins prioritized from the serial-mediation. Color denotes the dominant mechanism --- amplifying-dominant (red; > 70% of the protein’s pathways amplifying), compensatory-dominant (blue), or mixed (grey).

**Table 3.** Core mediating proteins. The 33 broadly-acting core proteins prioritized from the serial-mediation pathways (75th-percentile on all four breadth/effect-size metrics; cf. Figure 4), with dominant mechanism, number of significant pathways, metabolite classes spanned, max/mean absolute proportion mediated, and % of pathways amplifying.

| Protein | Dominant Mechanism | #path ways | #metabolite classes | Max prop mediated | Mean prop mediated | Percentage of amplifying |
| --- | --- | --- | --- | --- | --- | --- |
| APOF | Amplifying | 49 | 14 | 0.183 | 0.100 | 100.0 |
| APOD | Amplifying | 40 | 14 | 0.158 | 0.108 | 100.0 |
| APOM | Amplifying | 48 | 13 | 0.152 | 0.084 | 100.0 |
| MXRA8 | Amplifying | 42 | 12 | 0.137 | 0.087 | 88.1 |
| APOA1 | Amplifying | 35 | 12 | 0.128 | 0.083 | 100.0 |
| RTN4R | Amplifying | 38 | 13 | 0.110 | 0.078 | 92.1 |
| LDLR | Compensatory | 84 | 16 | 0.310 | 0.163 | 17.9 |
| SSC4D | Compensatory | 49 | 13 | 0.262 | 0.129 | 28.6 |
| **IGFBP2** | Compensatory | 50 | 12 | 0.251 | 0.120 | 24.0 |
| **LCAT** | Compensatory | 69 | 15 | 0.191 | 0.120 | 20.3 |
| **MFGE8** | Compensatory | 52 | 12 | 0.166 | 0.089 | 9.6 |
| **GHR** | Compensatory | 52 | 13 | 0.155 | 0.096 | 26.9 |
| **IGSF9** | Compensatory | 49 | 14 | 0.150 | 0.091 | 26.5 |
| **CES1** | Compensatory | 47 | 13 | 0.147 | 0.089 | 25.5 |
| **AFM** | Compensatory | 41 | 13 | 0.147 | 0.089 | 2.4 |
| **NPY** | Compensatory | 42 | 12 | 0.142 | 0.088 | 0.0 |
| **ERBB2** | Compensatory | 55 | 13 | 0.136 | 0.084 | 21.8 |
| **LAMP2** | Compensatory | 46 | 12 | 0.132 | 0.082 | 4.3 |
| **SSC5D** | Compensatory | 58 | 13 | 0.127 | 0.083 | 17.2 |
| **CNTN3** | Compensatory | 36 | 11 | 0.124 | 0.079 | 13.9 |
| **BCHE** | Compensatory | 44 | 12 | 0.102 | 0.071 | 29.5 |
| **ADIPOQ** | Mixed | 69 | 16 | 0.250 | 0.126 | 56.5 |
| **PLTP** | Mixed | 51 | 13 | 0.184 | 0.092 | 64.7 |
| **INHBC** | Mixed | 54 | 13 | 0.155 | 0.094 | 42.6 |
| **CPM** | Mixed | 51 | 12 | 0.146 | 0.087 | 31.4 |
| **PON3** | Mixed | 67 | 14 | 0.143 | 0.088 | 55.2 |
| **LPL** | Mixed | 41 | 12 | 0.125 | 0.089 | 46.3 |
| **PRAP1** | Mixed | 65 | 15 | 0.121 | 0.077 | 36.9 |
| **GSTA1** | Mixed | 40 | 11 | 0.113 | 0.078 | 40.0 |
| **IGFBP1** | Mixed | 52 | 15 | 0.110 | 0.074 | 44.2 |
| **PLAT** | Mixed | 44 | 11 | 0.109 | 0.078 | 36.4 |
| **SERPINF1** | Mixed | 39 | 11 | 0.108 | 0.071 | 41.0 |
| **FURIN** | Mixed | 45 | 11 | 0.102 | 0.075 | 35.6 |

### 3.7 Core protein mediation through large HDL subclasses

Across the 3,669 pathways, lipoprotein-particle metabolites dominated, and HDL was the single most represented metabolite class (1,019 pathways), ahead of very-low-density lipoprotein (VLDL, 751) and low-density lipoprotein (LDL, 371). To localize this signal, we focused on the 33 core proteins and the two largest HDL subclasses --- large HDL (average particle diameter 12.1 nm) and very large HDL (14.3 nm), 14 lipoprotein measures in total. 30 of the 33 core proteins and all 14 HDL-subclass metabolites participated in at least one core pathway, forming 306 core-protein × HDL-subclass pathways (150 through L-HDL, 156 through XL-HDL; Figure 5; Supplementary Table S6).

**Figure 5.**
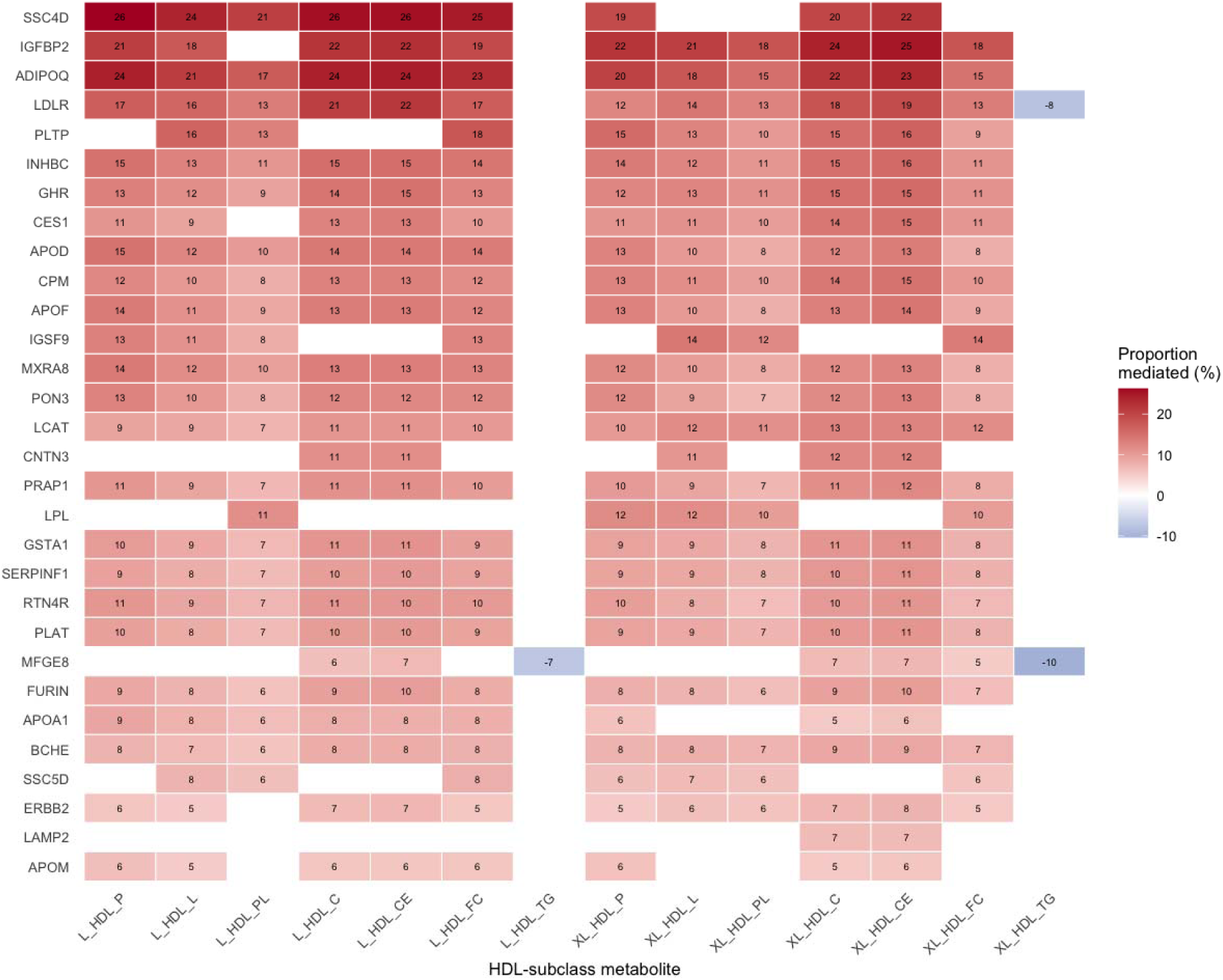
Core proteins mediate the IR-PRS -> CHD effect through HDL particle subclasses. Serial-mediation proportion (IR PRS -> core protein -> HDL metabolite -> CHD) for the 30 core proteins with ≥1 significant HDL-subclass pathway (rows) across 14 L-HDL and XL-HDL measures (columns). The value in each cell represents the proportion of the total effect mediated (×100), and the color indicates direction (red, amplifying; blue, compensatory). Lipoprotein-measure suffixes (Nightingale NMR): C, total cholesterol; CE, cholesteryl esters; FC, free cholesterol; PL, phospholipids; P, particle concentration; L, total lipids; SIZE, mean particle diameter. Subclass prefixes: XL, very large; L, large; M, medium; S, small.

The direction of these pathways was strikingly consistent: 303 of 306 (99.0%) were amplifying --- higher genetic IR was associated with a core-protein change that in turn tracked an HDL-subclass change associated with higher CHD risk --- with only three compensatory. The signal localized to the concentration and cholesterol/cholesteryl-ester content of X-HDL and XL-HDL particles rather than to bulk HDL-cholesterol or apolipoprotein summaries, and the amplifying core proteins were led by the HDL apolipoproteins themselves (APOA1, APOM, APOD, APOF). Together, these results identify large HDL particle subclasses as the dominant lipoprotein route through which the genetic component of insulin resistance is transmitted to CHD.

## 4. Discussion

In this study we combined multi-trait genetic discovery with large-scale proteomic and metabolomic mediation to dissect the molecular link between insulin resistance and coronary heart disease. By jointly modelling the share genetic signal of three IR-related metabolic traits using GenomicSEM, we defined a single latent IR factor, derived a multivariate GWAS of that factor, and used it to build a polygenic risk score that is associated with CHD in an independent UK Biobank European cohort. Layering causal-mediation and serial-mediation analyses on top of this genetic instrument, we found that the path from genetic insulin resistance to coronary risk runs predominantly through circulating proteins associated with HDL lipoprotein-particle subclasses --- specifically the L-HDL and XL-HDL --- rather than through conventional lipid measures such as total cholesterol, LDL cholesterol, or apolipoprotein.

The convergence of the mediation signals on HDL particle subclasses represents the central finding of this study. Insulin resistance has traditionally been associated with an atherogenic dyslipidemia characterized by low HDL cholesterol, elevated triglycerides, and a shift toward small, dense lipoprotein particles^30,31^. Our results refine this picture by localizing the genetic IR->CHD signal not simply to total HDL-C levels, but to specific alterations in HDL particle composition and subclass distribution, particularly L-HDL and XL-HDL particles. That 30 of the 33 broadly-acting core proteins and all 14 HDL-subclass metabolites participated in the 306 core-protein-HDL-subclass pathways, and that the overwhelming majority of these pathways (99.0%) were amplifying, indicates a coherent directional cascade in which genetically predicted insulin resistance perturbs a protein layer that in turn reshapes HDL-subclass biology associated with higher coronary risk. The smaller set of compensatory pathways suggests that some of this protein response is homeostatic, a nuance that single-mediator analyses would have missed.A central and initially counterintuitive feature of our results is the direction of the HDL signal: higher genetic insulin resistance was transmitted to greater CHD risk through increases in L-HDL and XL-HDL measures, so that these classically "protective" particles behaved as risk-amplifying mediators. This apparent paradox is consistent with a substantial body of genetic, epidemiological, and pharmacological evidence that HDL cholesterol is not causally cardioprotective. Mendelian-randomization analyses show that alleles raising HDL-C do not lower myocardial-infarction risk^32^, and a loss-of-function variant in the HDL receptor *SCARB1* that raises HDL-C nonetheless increases CHD risk^33^ --- demonstrating that impaired HDL clearance, and the resulting cholesterol-enriched, larger particles, can be harmful rather than protective. Pharmacological HDL-C raising through cholesteryl ester transfer protein inhibition has likewise failed to reduce, and in one trial increased, cardiovascular events^34,35^. Most directly, in people with type 2 diabetes, NMR-measured XL-HDL particles have been positively associated with mortality^36^, and large HDL particles confer risk at very high concentrations once apolipoprotein levels are accounted for^37^. In the insulin-resistant state, chronic glycoxidative modification renders HDL "dysfunctional," impairing its antioxidant, anti-inflammatory, and cholesterol-efflux capacity^38^. Our finding that the genetic component of insulin resistance reaches CHD specifically through the concentration and cholesterol content of L-HDL and XL-HDL particles fits this reframing: the risk localizes not to HDL-C per se, but to a shift in HDL particle biology that, in the setting of insulin resistance, tracks higher coronary risk.

Methodologically, our approach illustrates the value of treating insulin resistance as a latent genetic construct rather than relying on any single indicator. Because IR is not directly measurable from a single assay, a multi-trait factor model captures the shared biology while down-weighting trait-specific noise, and the resulting PRS provides an exposure instrument that is less prone to the attenuation and confounding that affect observational IR phenotypes. Embedding this instrument in a serial-mediation framework - rather than testing proteins and metabolites in isolation - allowed us to recover the sequential dependencies, i.e, from protein to metabolite, that are biologically expected but analytically invisible to single-mediator screens.

From a translational standpoint, the protein-HDL link identified here defines a candidate space for biomarkers and intervention. The core mediating proteins, prioritised for both the breadth and the magnitude of their pathway involvement, represent hypotheses for mechanistic follow-up and, potentially, for pharmacological modulation upstream of HDL-subclass remodelling. Besides, the finding that HDL particle subclasses rather than bulk HDL-C carry the mediating signal cautions against using total HDL-C as a surrogate endpoint and argues for subclass-resolved lipoprotein profiling in studies of insulin resistance and coronary risk.

This study has a few limitations. First, mediation analysis rests on no-unmeasured-confounding and correct-ordering assumptions; although we used a genetic exposure and adjusted for a broad covariate set, residual confounding (including by pleiotropy of the IR variants) and reverse causation between proteins and metabolites cannot be fully excluded^39^, and the serial ordering protein->metabolite is a modelling choice rather than an established causal sequence. Second, the analyses were conducted in a single European-ancestry cohort, so generalisability to other ancestries is untested and the PRS is expected to transfer imperfectly^40^. Third, the proteomic and metabolomic assays cover only a subset of the circulating proteome and metabolome (2,923 Olink proteins and 168 NMR biomarkers), and the differential sample coverage of the two platforms required different effective-sample-size thresholds, which may bias the relative representation of protein-versus metabolite-mediated pathways. Fourth, the one-factor model for the three IR indicators is just-identified, pro-insulin loads comparatively weakly on the latent factor, and a mild Heywood case was observed for fasting insulin; the factor should therefore be interpreted as a pragmatic summary of shared IR genetics rather than a definitive latent structure. Finally, CHD was defined from combined self-report and hospital records, and outcome misclassification would bias mediation estimates toward the null.

In summary, by integrating multi-trait genetic discovery with proteomic and metabolomic mediation, we map a proteome-to-HDL-subclass pathway that links the genetic component of insulin resistance to coronary heart disease. These findings nominate HDL lipoprotein-subclass biology, and the proteins that shape it, as a priority space for mechanistic, biomarker, and therapeutic investigation, and they demonstrate a generalizable framework for moving from genetic susceptibility to interpretable molecular intermediaries of disease.

## Data Availability

The IR-traits GWAS summary statistics analyzed here are publicly available: fasting insulin (GWAS Catalog accession GCST90002238), the Modified Stumvoll insulin sensitivity index (GCST90267573), and pro-insulin (MAGIC consortium, Broadaway et al., 2023, Am J Hum Genet, DOI: 10.1016/j.ajhg.2023.01.002). Individual-level UK Biobank phenotypic, genotypic, proteomic (Olink), and metabolomic (Nightingale NMR) data are available to approved researchers through the UK Biobank Access Management System (https://www.ukbiobank.ac.uk/).
The multivariate insulin-resistance GWAS summary statistics generated in this study, and the PRS-CS posterior SNP effect weights used to construct the polygenic score, are available at Synapse (project syn76570576) from the date of publication. Because both were derived solely from publicly available non-UK Biobank GWAS summary statistics, they contain no individual-level or UK Biobank-derived data and carry no redistribution restrictions.
Analysis code is available at https://github.com/RuiDongDR/ir-chd-mediation.

## Contributors

Rui Dong: Conceptualisation, Methodology, Software, Formal analysis, Data curation, Visualisation, Funding acquisition, Writing – original draft, Writing – review & editing. Jiao Fu: Conceptualisation, Validation, Investigation, Funding acquisition, Writing – original draft, Writing – review & editing. Bowen Zhang: Validation, Writing – review & editing. Qingling Zhang: Validation, Writing – review & editing. Yang Yan: Conceptualisation, Supervision, Project administration, Writing – review & editing. Guoliang Li: Conceptualisation, Supervision, Project administration, Funding acquisition, Writing – review & editing. Rui Dong and Jiao Fu contributed equally to this work. All authors have directly accessed and verified the underlying data reported in the manuscript. All authors read and approved the final version of the manuscript.

## Acknowledgements

This research was supported by institutional start-up funding from the Hetao Institute of Mathematics and Interdisciplinary Sciences (HIMIS) to R.D., the Clinical Research Award of the First Affiliated Hospital of Xi’an Jiaotong University, China (No.XJTU1AF2021CRF-001) to Guoliang Li, the National Natural Science Foundation of China (Grant No. 82571015) to Jiao Fu.

We thank the UK Biobank participants and the UK Biobank team. We also thank Prof. Gao Wang from Columbia University Irving Medical Center for helpful discussions and valuable feedback on this work.

## Data sharing statement

The IR-traits GWAS summary statistics analyzed here are publicly available: fasting insulin (GWAS Catalog accession GCST90002238), the Modified Stumvoll insulin sensitivity index (GCST90267573), and pro-insulin (MAGIC consortium, Broadaway et al., 2023, Am J Hum Genet, DOI: 10.1016/j.ajhg.2023.01.002). Individual-level UK Biobank phenotypic, genotypic, proteomic (Olink), and metabolomic (Nightingale NMR) data are available to approved researchers through the UK Biobank Access Management System (https://www.ukbiobank.ac.uk/).

The multivariate insulin-resistance GWAS summary statistics generated in this study, and the PRS-CS posterior SNP effect weights used to construct the polygenic score, are available at Synapse (project syn76570576) from the date of publication. Because both were derived solely from publicly available non-UK Biobank GWAS summary statistics, they contain no individual-level or UK Biobank-derived data and carry no redistribution restrictions.

Analysis code is available at https://github.com/RuiDongDR/ir-chd-mediation.

## Abbreviations

ACME: average causal mediation effect;
BMI: body mass index;
CHD: coronary heart disease;
CI: confidence interval;
DWLS: diagonally weighted least squares;
FDR: false discovery rate;
GenomicSEM: Genomic Structural Equation Modelling;
GWAS: genome-wide association study;
HDL: high-density lipoprotein;
ICD: International Classification of Diseases;
INFO: imputation information score;
IR: insulin resistance;
ISI: (Modified Stumvoll) insulin sensitivity index;
LD: linkage disequilibrium;
LDL: low-density lipoprotein;
LDSC: LD Score Regression;
L-HDL: large high-density lipoprotein;
MAF: minor allele frequency;
MET: metabolic equivalent of task;
NMR: nuclear magnetic resonance;
NPX: normalized protein expression;
NT-proBNP: N-terminal pro-B-type natriuretic peptide;
OR: odds ratio;
PRS: polygenic risk score;
PRS-CS: polygenic risk score - continuous shrinkage;
QC: quality control;
Q_SNP_: per-SNP heterogeneity statistic;
SD: standard deviation;
SE: standard error;
SEM: structural equation model;
SNP: single-nucleotide polymorphism;
T2DM: type 2 diabetes mellitus;
UKB-PPP: UK Biobank Pharma Proteomics Project;
VLDL: very-low-density lipoprotein;
XL-HDL: very large high-density lipoprotein;
λ_GC_: genomic-control inflation factor. Lipoprotein-measure suffixes in Nightingale NMR:
C: total cholesterol;
CE: cholesteryl esters;
FC: free cholesterol;
PL: phospholipids;
P: particle concentration;
L: total lipids;
SIZE: mean particle diameter. Subclass prefixes:
XL: very large;
L: large;
M: medium;
S: small.

## Web resources

EasyQC, https://www.uni-regensburg.de/medizin/epidemiologie-praeventivmedizin/genetische-epidemiologie/software/ ; gwaslab, https://cloufield.github.io/gwaslab/ ; LD Score Regression (LDSC), https://github.com/bulik/ldsc ; GenomicSEM, https://github.com/GenomicSEM/GenomicSEM ; PRS-CS, https://github.com/getian107/PRScs ; lavaan, https://lavaan.ugent.be/ ; mediation (R package), https://cran.r-project.org/package=mediation ; ukbnmr (R package), https://cran.r-project.org/package=ukbnmr ; UK Biobank, https://www.ukbiobank.ac.uk/.

## References

1. Khan MA, Hashim MJ, Mustafa H, Baniyas MY, Al Suwaidi SKBM, AlKatheeri R, et al. Global epidemiology of ischemic heart disease: Results from the global burden of disease study. Cureus. 12(7):e9349. doi:10.7759/cureus.9349 PubMed PMID: 32742886; PubMed Central PMCID: PMC7384703.

2. Collaboration TERF. Diabetes mellitus, fasting blood glucose concentration, and risk of vascular disease: A collaborative meta-analysis of 102 prospective studies. The Lancet. 2010 Jun 26;375(9733):2215–22. doi:10.1016/S0140-6736(10)60484-9 PubMed PMID: 20609967.

3. Beverly JK, Budoff MJ. Atherosclerosis: Pathophysiology of insulin resistance, hyperglycemia, hyperlipidemia, and inflammation. Journal of Diabetes. 2020;12(2):102–4. doi:10.1111/1753-0407.12970

4. Wang Z, Chen J, Zhu L, Jiao S, Chen Y, Sun Y. Metabolic disorders and risk of cardiovascular diseases: A two-sample mendelian randomization study. BMC Cardiovasc Disord. 2023 Oct 31;23:529. doi:10.1186/s12872-023-03567-3 PubMed PMID: 37907844; PubMed Central PMCID: PMC10617200.

5. Bulik-Sullivan BK, Loh PR, Finucane HK, Ripke S, Yang J, Patterson N, et al. LD score regression distinguishes confounding from polygenicity in genome-wide association studies. Nat Genet. 2015 Mar;47(3):291–5. doi:10.1038/ng.3211

6. Bulik-Sullivan B, Finucane HK, Anttila V, Gusev A, Day FR, Loh PR, et al. An atlas of genetic correlations across human diseases and traits. Nat Genet. 2015 Nov;47(11):1236–41. doi:10.1038/ng.3406

7. Grotzinger AD, Rhemtulla M, de Vlaming R, Ritchie SJ, Mallard TT, Hill WD, et al. Genomic SEM provides insights into the multivariate genetic architecture of complex traits. Nat Hum Behav. 2019 May;3(5):513–25. doi:10.1038/s41562-019-0566-x PubMed PMID: 30962613; PubMed Central PMCID: PMC6520146.

8. Sun BB, Chiou J, Traylor M, Benner C, Hsu YH, Richardson TG, et al. Plasma proteomic associations with genetics and health in the UK biobank. Nature. 2023 Oct;622(7982):329–38. doi:10.1038/s41586-023-06592-6 PubMed PMID: 37794186; PubMed Central PMCID: PMC10567551.

9. Julkunen H, Cichońska A, Tiainen M, Koskela H, Nybo K, Mäkelä V, et al. Atlas of plasma NMR biomarkers for health and disease in 118,461 individuals from the UK biobank. Nat Commun. 2023 Feb 3;14(1):604. doi:10.1038/s41467-023-36231-7

10. Lotta LA, Gulati P, Day FR, Payne F, Ongen H, van de Bunt M, et al. Integrative genomic analysis implicates limited peripheral adipose storage capacity in the pathogenesis of human insulin resistance. Nat Genet. 2017 Jan;49(1):17–26. doi:10.1038/ng.3714 PubMed PMID: 27841877; PubMed Central PMCID: PMC5774584.

11. Ye C, Dou C, Liu D, Kong L, Chen M, Xu M, et al. Multivariate genome-wide analyses of insulin resistance unravel novel loci and therapeutic targets for cardiometabolic health. Nat Commun. 2025 Nov 17;16(1):10057. doi:10.1038/s41467-025-64985-9

12. Du J, Zhou M, Raffield LM, Zhou R, Li Y, Chen C, et al. Multi-omics integration predicts 17 disease incidences in the UK biobank. medRxiv: The Preprint Server for Health Sciences; 2025. doi:10.1101/2025.08.01.25332841 PubMed PMID: 40799968; PubMed Central PMCID: PMC12340880.

13. Li Y, Li D, Lin J, Zhou L, Yang W, Yin X, et al. Proteomic signatures of type 2 diabetes predict the incidence of coronary heart disease. Cardiovasc Diabetol. 2025 Mar 14;24(1):120. doi:10.1186/s12933-025-02670-3 PubMed PMID: 40087642; PubMed Central PMCID: PMC11909814.

14. Qian H, Wu C, Li B, Rosenzweig A, Wang M. Plasma proteomics linking primary and secondary diseases: Insights into molecular mediation from UK biobank data. medRxiv: The Preprint Server for Health Sciences; 2025. doi:10.1101/2025.08.29.25334726 PubMed PMID: 40950430; PubMed Central PMCID: PMC12424883.

15. Ge T, Chen CY, Ni Y, Feng YCA, Smoller JW. Polygenic prediction via bayesian regression and continuous shrinkage priors. Nat Commun. 2019 Apr 16;10(1):1776. doi:10.1038/s41467-019-09718-5

16. Chen J, Spracklen CN, Marenne G, Varshney A, Corbin LJ, Luan J, et al. The trans-ancestral genomic architecture of glycemic traits. Nat Genet. 2021 Jun;53(6):840–60. doi:10.1038/s41588-021-00852-9 PubMed PMID: 34059833; PubMed Central PMCID: PMC7610958.

17. Williamson A, Norris DM, Yin X, Broadaway KA, Moxley AH, Vadlamudi S, et al. Genome-wide association study and functional characterization identifies candidate genes for insulin-stimulated glucose uptake. Nat Genet. 2023 Jun;55(6):973–83. doi:10.1038/s41588-023-01408-9 PubMed PMID: 37291194; PubMed Central PMCID: PMC7614755.

18. Broadaway KA, Yin X, Williamson A, Parsons VA, Wilson EP, Moxley AH, et al. Loci for insulin processing and secretion provide insight into type 2 diabetes risk. Am J Hum Genet. 2023 Feb 2;110(2):284–99. doi:10.1016/j.ajhg.2023.01.002 PubMed PMID: 36693378; PubMed Central PMCID: PMC9943750.

19. Winkler TW, Day FR, Croteau-Chonka DC, Wood AR, Locke AE, Mägi R, et al. Quality control and conduct of genome-wide association meta-analyses. Nat Protoc. 2014 May;9(5):1192–212. doi:10.1038/nprot.2014.071 PubMed PMID: 24762786; PubMed Central PMCID: PMC4083217.

20. 1000 Genomes Project Consortium, Auton A, Brooks LD, Durbin RM, Garrison EP, Kang HM, et al. A global reference for human genetic variation. Nature. 2015 Oct 1;526(7571):68–74. doi:10.1038/nature15393 PubMed PMID: 26432245; PubMed Central PMCID: PMC4750478.

21. Park S, Kim S, Kim B, Kim DS, Kim J, Ahn Y, et al. Multivariate genomic analysis of 5 million people elucidates the genetic architecture of shared components of the metabolic syndrome. Nat Genet. 2024 Nov;56(11):2380–91. doi:10.1038/s41588-024-01933-1 PubMed PMID: 39349817; PubMed Central PMCID: PMC11549047.

22. He Y, Koido M, Shimmori Y, Kamatani Y. GWASLab: A python package for processing and visualizing GWAS summary statistics [Internet]. 2023 May 1. doi:10.51094/jxiv.370

23. Sherry ST, Ward MH, Kholodov M, Baker J, Phan L, Smigielski EM, et al. dbSNP: The NCBI database of genetic variation. Nucleic Acids Res. 2001 Jan 1;29(1):308–11. doi:10.1093/nar/29.1.308 PubMed PMID: 11125122; PubMed Central PMCID: PMC29783.

24. O’Leary NA, Wright MW, Brister JR, Ciufo S, Haddad D, McVeigh R, et al. Reference sequence (RefSeq) database at NCBI: Current status, taxonomic expansion, and functional annotation. Nucleic Acids Res. 2016 Jan 4;44(D1):D733–45. doi:10.1093/nar/gkv1189

25. Ge T, Chen CY, Ni Y, Feng YCA, Smoller JW. Polygenic prediction via bayesian regression and continuous shrinkage priors. Nat Commun. 2019 Apr 16;10(1):1776. doi:10.1038/s41467-019-09718-5

26. Ritchie SC, Surendran P, Karthikeyan S, Lambert SA, Bolton T, Pennells L, et al. Quality control and removal of technical variation of NMR metabolic biomarker data in ∼120,000 UK biobank participants. Sci Data. 2023 Jan 31;10(1):64. doi:10.1038/s41597-023-01949-y

27. Imai K, Keele L, Tingley D. A general approach to causal mediation analysis. Psychol Methods. 2010 Dec;15(4):309–34. doi:10.1037/a0020761 PubMed PMID: 20954780.

28. Rosseel Y. lavaan: An R package for structural equation modeling. Journal of Statistical Software [Internet]. 2012 [cited 2026 Jul 31];048(i02). Available from: https://ideas.repec.org//a/jss/jstsof/v048i02.html

29. Benjamini Y, Hochberg Y. Controlling the false discovery rate: A practical and powerful approach to multiple testing. Royal Statistical Society Journal Series B: Methodological. 1995 Jan 1;57(1):289–300. doi:10.1111/j.2517-6161.1995.tb02031.x

30. Howard BV. Insulin resistance and lipid metabolism. The American Journal of Cardiology. 1999 Jul 8;84(1, Supplement 1):28–32. doi:10.1016/S0002-9149(99)00355-0

31. Hirano T. Pathophysiology of diabetic dyslipidemia. Journal of Atherosclerosis and Thrombosis. 2018;25(9):771–82. doi:10.5551/jat.RV17023

32. Voight BF, Peloso GM, Orho-Melander M, Frikke-Schmidt R, Barbalic M, Jensen MK, et al. Plasma HDL cholesterol and risk of myocardial infarction: A mendelian randomisation study. The Lancet. 2012 Aug 11;380(9841):572–80. doi:10.1016/S0140-6736(12)60312-2 PubMed PMID: 22607825.

33. Zanoni P, Khetarpal SA, Larach DB, Hancock-Cerutti WF, Millar JS, Cuchel M, et al. Rare variant in scavenger receptor BI raises HDL cholesterol and increases risk of coronary heart disease. Science. 2016 Mar 11;351(6278):1166–71. doi:10.1126/science.aad3517 PubMed PMID: 26965621; PubMed Central PMCID: PMC4889017.

34. Barter PJ, Caulfield M, Eriksson M, Grundy SM, Kastelein JJP, Komajda M, et al. Effects of torcetrapib in patients at high risk for coronary events. N Engl J Med. 2007 Nov 22;357(21):2109–22. doi:10.1056/NEJMoa0706628 PubMed PMID: 17984165.

35. Lincoff AM, Nicholls SJ, Riesmeyer JS, Barter PJ, Brewer HB, Fox KAA, et al. Evacetrapib and cardiovascular outcomes in high-risk vascular disease. N Engl J Med. 2017 May 18;376(20):1933–42. doi:10.1056/NEJMoa1609581 PubMed PMID: 28514624.

36. Jin Q, Lau ESH, Luk AO, Tam CHT, Ozaki R, Lim CKP, et al. High-density lipoprotein subclasses and cardiovascular disease and mortality in type 2 diabetes: Analysis from the hong kong diabetes biobank. Cardiovasc Diabetol. 2022 Dec 31;21(1):293. doi:10.1186/s12933-022-01726-y PubMed PMID: 36587202; PubMed Central PMCID: PMC9805680.

37. van der Steeg WA, Holme I, Boekholdt SM, Larsen ML, Lindahl C, Stroes ESG, et al. High-density lipoprotein cholesterol, high-density lipoprotein particle size, and apolipoprotein a-I: Significance for cardiovascular risk: the IDEAL and EPIC-norfolk studies. J Am Coll Cardiol. 2008 Feb 12;51(6):634–42. doi:10.1016/j.jacc.2007.09.060 PubMed PMID: 18261682.

38. Bonilha I, Zimetti F, Zanotti I, Papotti B, Sposito AC. Dysfunctional high-density lipoproteins in type 2 diabetes mellitus: Molecular mechanisms and therapeutic implications. J Clin Med. 2021 May 21;10(11):2233. doi:10.3390/jcm10112233 PubMed PMID: 34063950; PubMed Central PMCID: PMC8196572.

39. Böhnke JR. Explanation in causal inference: Methods for mediation and interaction. Q J Exp Psychol (Hove). 2016 Jun;69(6):1243–4. doi:10.1080/17470218.2015.1115884 PubMed PMID: 28190383.

40. Privé F, Aschard H, Carmi S, Folkersen L, Hoggart C, O’Reilly PF, et al. Portability of 245 polygenic scores when derived from the UK biobank and applied to 9 ancestry groups from the same cohort. Am J Hum Genet. 2022 Jan 6;109(1):12–23. doi:10.1016/j.ajhg.2021.11.008 PubMed PMID: 34995502; PubMed Central PMCID: PMC8764121.

