## Supplementary figures and images for "Dissecting the Genetic Link Between Insulin Resistance and Coronary Heart Disease: A Multi-Trait GWAS and Proteomic-Metabolomic Mediation Analysis"

### Supplementary Figure S1

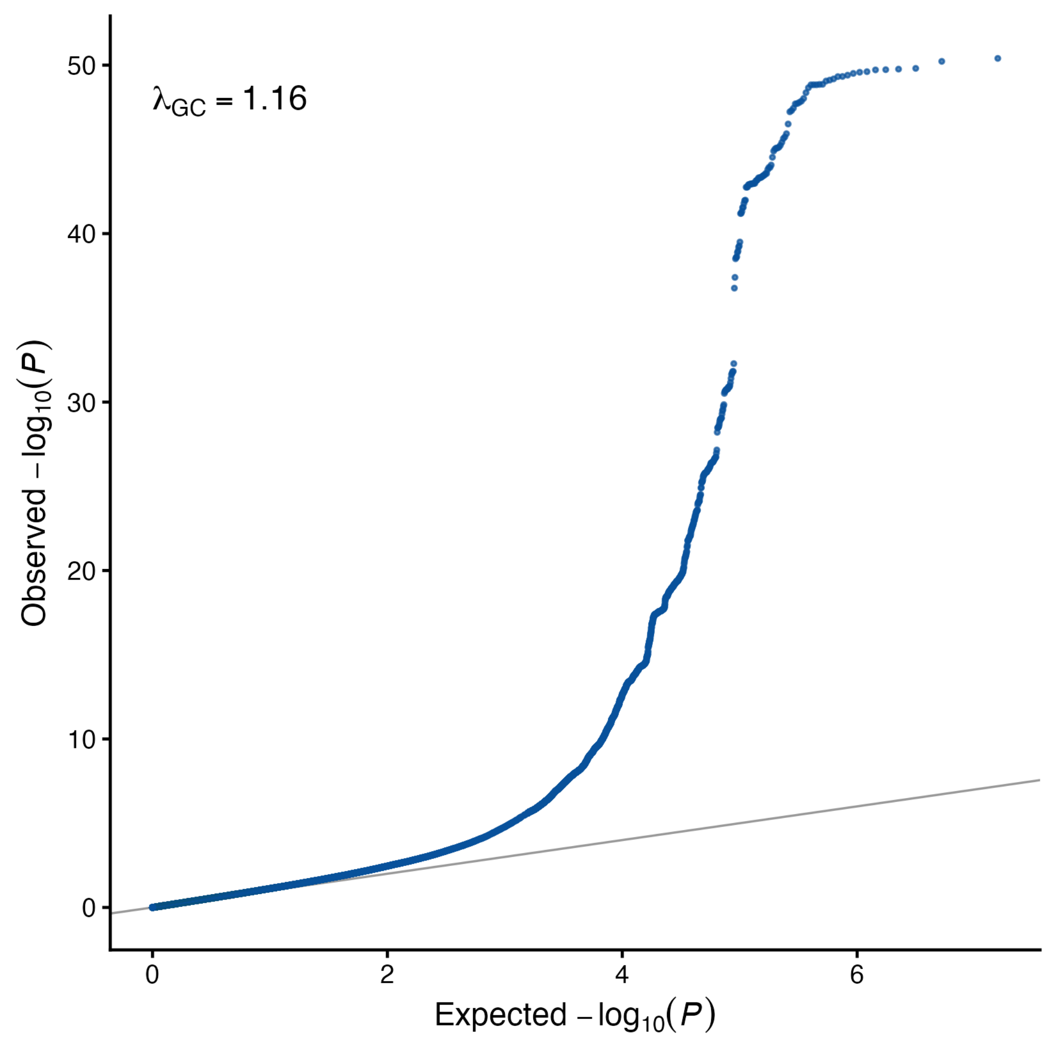
